# Determinants of the use of Digital One Health amongst medical and veterinary students at the University of Buea, Cameroon

**DOI:** 10.1101/2025.04.16.25325946

**Authors:** Elijah Forteh, Moses Njutain Ngemenya, Ginyu Innocertia Kwalar, Eleanor Ngeh-Nkeng Alembong, Elvis Asangbeng Tanue, Odette Dzemo Kibu, Jude Dzevela Kong, Dickson Shey Nsagha

**Affiliations:** Department of Public Health and Hygiene, Faculty of Health Sciences, University of Buea, Buea, Cameroon; Department of Medical Laboratory Sciences, Faculty of Public Health, University of Buea, Cameroon; Department of Mathematics & Statistics, York University, Toronto, Canada

**Author notes:** Corresponding author: Professor Dickson Shey Nsagha.

**Keywords:** One Health, Digital health, Digital One Health, medical and veterinary students, Cameroon

## Abstract

Digital tools help foster collaboration within the One Health (OH) framework. Identifying factors that influence the use of Digital One Health (DOH) among medical and veterinary students is critical to improving its inclusion into future healthcare practices, hence the need of this study.

This is a school-based cross-sectional study was carried out from December 2023 to June 2024. Data collection on socio-demography, awareness of DOH, capacity building, and digital competencies were collected as from 19^th^ February to 5^th^ May 2024. Descriptive statistics used to summarize awareness, capacity building, and digital competencies of participants. Multiple logistic regression helped identify determinants of the use of DOH.

The results revealed that, of 400 participants enrolled, 318 (79.5%) had not heard of DOH, 334 (83.4%) had never received capacity building on DOH. Encountering DOH information from health information sources (AOR= 3.88 [95%CI: 1.66 – 9.03], p=0.002), receiving capacity building on DOH (AOR= 3.06 [95%CI: 1.15 – 8.16], p=0.026), having data privacy and security concerns (AOR= 3.75 [95%CI: 1.45 – 9.73], p=0.007), knowledge of five digital technologies used in OH (AOR= 15.43 [95%CI: 3.69 – 64.52], p=0.001) and access to at least one school technological infrastructure (AOR= 2.94 [95%CI: 1.23 – 7.03], p=0.015) significantly increase the odds of DOH engagement.

In Conclusion, Awareness and capacity building on DOH is low amongst medical and veterinary students of the University of Buea. Encountering DOH information from health information sources, capacity building, data privacy/ security concerns, knowledge of the various digital technologies used in OH, and access to school technological infrastructures, is vital to DOH engagement by medical and veterinary students at the University of Buea.

## Introduction

One Health (OH) is an umbrella concept that encompasses all disciplines that broadly deal with the health of humans, animals, and their surrounding environment [1]. The modern origins of OH date back to 2004. One Health was introduced as part of the 12 Manhattan Principles, which called for an international, interdisciplinary approach for preventing diseases specifically animal-human transmissible and communicable diseases [2]. Therefore, systematic perspectives on life sciences and the environment were brought together to design and implement programs, policies, and regulations for achieving better public health outcomes. The World Health Organization (WHO) has associated OH with sustainable development goals [3,4]. One Health also involves the evaluation and monitoring of the impact of environmental hazards on health care systems, public health, biodiversity, and food security [1]. In summary, OH encompasses the interconnections between humans, animals, plants, and the global ecological environment [5,8]. A major component of OH has been data, with one of the main concerns being the actual and prospective spread of diseases among populations of domestic animals, wildlife, and humans. The current understanding of OH, which is a global strategy centered on population growth, industrialization, and geopolitical challenges, emphasizes the importance of data in evaluating health risks associated with the emergence or reemergence of infectious or non-infectious diseases through a transdisciplinary, holistic approach that emphasizes the dynamics between humans, animals, and ecosystems [9].

The Digital One Health (DOH) framework, published in 2021, emphasizes the digital linkages between human health, animal health, plant health, and the environment [10]. The DOH framework involves using digital capabilities in collecting, processing, and sharing data, information, and knowledge across multisectoral domains in the OH space. Surveillance of human and animal pathogens and genomes by public health professionals, healthcare practitioners, veterinarians and citizen scientists will inform an ecosystem approach to disease management, prevention, and future planning [10]. Citizen science initiatives using digital sensors and other digitized data collection techniques have contributed to the amassing of databases about phenomena such as the environmental effects of pollution, localized climate change impacts, loss of species diversity and efforts to recover cleared land [11]. DOH provides innovative options for integrating human, veterinary, and environmental data to support individualized therapies, ensuring “predictive, personalized, preventive, participatory (P4)” processes from start to finish [12]. The evolution of digital technology has resulted in evidence-based, accessible, upper-level, and holistic methods that are capable of accelerating biomedical research and enhancing public health efficacy [1]. Reflecting upon multiple perspectives during the Coronavirus disease 2019 (COVID-19) pandemic can provide opportunities for revisiting and understanding the limitations of public health, human and veterinary health care, and environmental management systems [13,14]. DOH encompasses various technologies such as telemedicine, mobile applications, wearable devices, data analytics, and electronic health records that enable the collection, analysis, and sharing of health information to improve animal and human health outcomes [15]. It also includes the use of artificial intelligence and machine learning algorithms to support decision-making and diagnosis in veterinary and human medicine. Despite the potential benefits of DOH, its implementation and adoption face several challenges. These challenges include limited interoperability between different platforms and systems, concerns regarding data privacy and security, regulatory barriers, and resistance from veterinary professionals to embrace digital technologies [16]. Digital health has been shown to improve the efficiency and scale of health service delivery in resource-limited settings, such as Cameroon [17]. With the right investments and policies, digital health has the potential to significantly enhance healthcare access and quality in Cameroon [18-20].

The government of Cameroon has also begun to prioritize digital health as part of its national healthcare agenda. Initiatives such as the development of a national electronic health(e-health) strategy and the establishment of a national e-health agency demonstrate the commitment to advancing digital healthcare in the country [21].

However, despite substantial breakthroughs in new digital technologies and processes, the slow adoption of e-health remains a challenge [22]. There has been insufficient capacity building among local and regional players [23]. Without major investment in training, the adoption of digital tools will be sluggish. Health workers have an important surveillance role in detecting illness outbreaks and their propagation, and when equipped with digital tools, they can report this information quickly [24]. In Cameroon, the Ministry of Public Health (MOH) released the National Digital Health Strategic Plan (NDHSP) 2020-2024, which stated that most health staff and users, especially those in rural areas, do not have digital skills, with many doctors and nurses primarily engaged in their technical work and consider Information and Communication Technology (ICT) to be an additional burden that takes them away from their main tasks [21]. Also, the lack of knowledge on OH by medical and veterinary students has been reported by different authors [25-27]. Hence this study was aimed at assessing the factors that influence the utilization of DOH amongst medical and veterinary students at the University of Buea.

## Methods

### Study design and settings

A school-based cross-sectional study was conducted in the University of Buea from December 2023 to June 2024. The University of Buea is an Anglo-Saxon university in the South-West region of Cameroon with four campuses located in Molyko, Clerks-quarter, Bomaka and Kumba. Overall, the University of Buea is made up of eight faculties which includes: Faculty of Arts, Science, Health Sciences, Law and Political Science, Social and Management Sciences, Agriculture and Veterinary Medicine, Education, and Engineering and Technology; and three schools, which are the Advanced school of Translators and Interpreters, College of Technology, and Higher Technical Teachers Training College. The medicine program is under the faculty of health sciences, in the campus at Bomaka-Buea which is a program that runs for six years plus one research year. The veterinary medicine program on the other hand is under the faculty of veterinary medicine and agriculture, located in the main campus (Molyko-Buea) and is a six-year program.

### Study population and sampling

This study targeted undergraduate students at the University of Buea who gave consent to participate. We included medical and veterinary students who gave consent and excluded students who were sick and/or unavailable at the time of the study.

Using Cochran’s formula [28], and an expected proportion of 50% due to limited literature, we obtained a minimum sample size of 385. All the program levels were taken into consideration and study participants were selected using convenience sampling.

### Ethical consideration

Ethical approval was obtained from the Institutional Review Board of the Faculty of Health Sciences University of Buea (reference number: 2024/2321-01/UB/SG/IRB/FHS) followed by authorization to carry out research from the Regional Delegation of Public Health for the Southwest (P42/MINSANTE/SWR/RDPH/CB.PT/578/678), and the Registrar of the University of Buea (2024/1327/UB/REG/ARD) Informed consent was obtained from all participants.

### Data collection and analysis

Structured questionnaires used to collect data on socio-demography, awareness of DOH, capacity building, and the digital competencies. The questionnaire was self-administered in the English language. Data collection took place from the 19^th^ of February 2024 to 05^th^ of May 2024 Data analysis was done using Statistical Package for Social Sciences (SPSS) version 25 (SPSS, IBM, Chicago, U.S.A). Descriptive statistics was used to summarize the awareness, capacity building, and digital competencies of participants, and multiple logistic regression used to identify determinants of the use of DOH.

## Results

### Socio-demographic characteristics of the study participants

The 400 participants ages ranged 16years to 29years, a mean age of 21.7 (SD 2.1). 201 (50.2) were males and 199 (49.8) females. 317 (79.3%) were medical students and 83 (20.7%) veterinary students.

### Awareness of Digital One Health

Out of 400 students 226 (96.5%) reported that digital services were relevant. 318 (79.5%) reported they had not heard of DOH. Rating their level of understanding of integration of digital technologies in OH, out of 380 responses, a majority of 171 (45%) participants reported to have partial understanding. 270 (70.1%) participants reported health care providers as health information source and a majority of 290 (72.6%) had not encountered DOH information source(s). Most participants reported to know mobile applications (51%), and telehealth/telemedicine (49.4%) as DOH components (table 1).

### Capacity building on Digital One Health

Majority of 334 (83.4%) participants reported to have never received training on DOH and of those who had received, 35 (52.5%) reported it being through lectures/academic settings, while 11 (17.5%), and 20 (30%) through professional development/training programs and seminar/conference respectively (table 2).

### Digital competencies of participants

Of 400 participants, 380 (95%) had received ICT classes and 151 (51%) reported to be moderately confident in using digital services. 184 (46%) participants were somewhat familiar with digital online health information retrieval. 232 (58%) reported moderate access to digital health technologies. 229 (59.2%) participants had access to one school technology, and the most accessed school technology the school IT centre (52.7%). 188 (46.9%) participants had access to three technologies at home. 321 (80.3%) participants reported that schoolwork load did not prevent DOH services usage, and 219 (54.8%) reported having sufficient personal time for its use. 276 (69%) reported data privacy and security concerns with DOH services (table 3).

### Awareness, capacity building, and digital competencies of participants based on program

More Veterinary students (98%) reported familiarity with the OH concept, while 49.7% of medical students reported to not have any familiarity with the OH concept. Most of the veterinary students (72.1%) reported all three components (animal health, human health, and environmental health), compared to 56% of medical students knowing all three. All veterinary students reported that digital services were relevant in OH. More medical students (70.8%) had never heard of DOH. Most of the veterinary students (60.9%) reported to have partial understanding of integration of digital technologies in OH while majority of the medical students reported having limited understanding (47%). More medical students (64.6%) had encountered DOH information from their source(s) (healthcare providers, internet/social media, government health agencies, and television and radio) compared to 51% for veterinary students. More veterinary medicine students (44%) had received capacity building on DOH compared to 12% of medical students, and most veterinary student (75.5%) reported to have engagement with DOH compared to 68.4% of medical students. Both medical (97.9%) and veterinary (100%) students reported being confident in using digital devices (computers, smartphones, and tablets), and were both familiar with digital online health information retrieval. 57.4% and 60% of medical and veterinary students respectively, reported to have moderate access to digital health technologies/resources. A majority of medical (53.7%) and veterinary (82.6%) students reported to have access to at least one of the school technological infrastructures (IT center and school Wi-Fi). A slight majority of medical (51.7%) and veterinary (66%) students reported having sufficient personal time to utilize DOH services. Most of the medical (69.8%) and veterinary (66%) students reported to having data privacy and security concerns with respect to using DOH services (table 4).

### Determinants of utilization of Digital One Health among participants

After controlling for confounders, the use of DOH was independently associated to; encountering DOH information from one’s health information sources, capacity building, number of school technologies you have access to, data privacy and security concerns and knowledge of the various digital technologies used in OH. The odds of a participant who encountered DOH information from their health information sources engaging with DOH services was 3.33 times higher than a participant who did not (AOR= 3.877 [95%CI: 1.664 – 9.034], p=0.002). Those who had received capacity building on DOH were 3.06 times more likely to use it than participants who had not (AOR= 3.059 [95%CI: 1.146 – 8.161], p=0.026). The odds of engaging with DOH services were 3.75 times higher for those with data privacy and security concerns compared to participants without concerns (AOR= 3.753 [95%CI: 1.447 – 9.73], p=0.007). Also, those who knew six digital technologies used in OH were 15.43 times more likely to use DOH services compared to those who knew only one (AOR= 15.427 [95%CI: 3.689 – 64.521], p=0.001). Lastly, the odds of a participant who had access to at least one school technological infrastructure using DOH services was 2.94 times higher than participants who had no access (AOR= 2.94 [95%CI: 1.229 – 7.031], p=0.015) (table 5).

## Discussion

We observed that more veterinary students (82%) were familiar with OH as compared to medical students (60.3%). Similarly, Terribadge et al in 2020 in Nigeria reported that veterinary students (65%) were generally more knowledgeable on OH than medical students (31.7%) [29]. In Nepal, Subedi et al in 2022 also reported that majority (55.6%) of veterinary medicine students had satisfactory knowledge scores on OH [30]. However, Franco-Martinez et al in Spain (2020) reported that just 20% of veterinary students were familiar with OH, with 80% having poor or very poor Knowledge on OH [31], of which this disparity might be due to the presence of confounding variables. Contrastingly too, Roopnarine et al in the Caribbean (2022) reported that majority (75%) of medical students were familiar with OH [32]. These disparities could be because of the different study settings.

On familiarity with OH, human health had the majority selection of 212 (88.3%) times similar to what Roopnarine et al reported that human health emerged as the key component associated with OH [32]. Furthermore, from our study, majority veterinary students reported all three OH components. Subedi et al also reported that majority (91.9%) of veterinary students reported that OH covers all three interconnections (human, animal, and environmental health) [33]. However, Franco-Martinez et al reported that majority of Veterinary students failed to include all three OH components [31]. These contrasting findings may be due to different study settings.

From our study, though a majority of participants reported that they had not heard of DOH, they reported that digital services were relevant in OH, which was the opinion of both medical and veterinary students. This, however, is different from the findings of Nguyen et al in Vietnam [34], who reported that medical students’ score of perceived benefits of e-health tools was low. This disparity can be because of the difference in knowledge of participants on digital technologies.

A majority (27.9%) of students reported being aware of at least one of six of the digital technologies used in OH and mobile applications identified the most and this may be due to the widespread use and integration of smartphones and mobile technology into daily life.

Our study also revealed that majority of participants had received Information and Communication Technology (ICT) classes which was consistent for both medical and veterinary students. However, Evbuomwan et al in Nigeria (2020), reported that not up to one-third of medical students had ever taken a course that is related to ICT [35]. This huge contrast may be due to disparities in their curricula.

Majority (92.2%) of participants had familiarity with digital online health information retrieval which is similar to what Asefeh-Asemi et al reported in Isfahan (2005) that majority (69.6%) of the students had familiarity with digital resources available to them in their academic libraries [36]. However, Martzoukou et al in Scotland (2024) reported low competencies in digital literacy dimensions like information literacy, digital research, digital innovation among students, all of which are imperative for their studies and for their future professional careers [37]. This low competency could be because students rely more on passive consumption like watching videos or browsing social media rather than engaging in activities that promote digital literacy like coding. From our study a low proportion of students had received capacity building on DOH and the majority came from veterinary students. Similarly, Sisira et al in Australia (2022) reported that most medical students had not received formal education and training on digital health [38]. This low proportion could be because medical as well as veterinary medicine curricula may not prioritize digital health technologies.

Our study revealed that, encountering DOH information from one’s health information sources, receiving capacity building on DOH, having data privacy and security concerns, knowledge of six digital technologies used in OH and having access to at least one school technology were the groups that were most likely to have engaged with DOH services. Borges et al in Denmark (2023), also reported capacity building (training / educational programs) to be one of the determinants of the use of digital health technologies by health care professionals, where they stated that offering training and educational activities increases the positive experience of participants [39]. Nguyen et al in Vietnam (2022), reported e-literacy to be one of the determinants of adoption of eHealth technologies among medical students. However, they also reported age and access to internet as determinants of adoption of eHealth technologies, which were not significant in our study [34]. On the other hand, Demsash et al in Ethiopia (2023) reported that adequate internet access, good knowledge, favourable attitudes, previous computer experience, and secondary / higher educational status of parents, were all determinants of digital technology utilization among health science students of which these variables were not significant in our study [40].

As Strength, our study highlights the level of awareness of students and the gaps in training on digital technologies in OH, and has identified the factors influencing the use and integration of digital technologies in the context of OH by medical and veterinary medicine students at the University of Buea.

## Conclusion

The low awareness of students on DOH might highlight a substantial lack of understanding among students regarding the value and potential applications of digital technologies within the OH framework. This low awareness may hinder their ability to effectively engage with digital tools for interdisciplinary collaboration and problem-solving. In addition, the level of capacity building of medical and veterinary students at the University of Buea on DOH was low and may impede their ability to effectively apply digital tools for complex health challenges. Lastly, factors such as technological proficiency, awareness of OH concepts, capacity building, interdisciplinary education, and institutional support play crucial roles in shaping students’ engagement with DOH. This study highlights the need for targeted interventions and educational initiatives at the University community to improve on students’ involvement in the domain of DOH. Educational institutions could invest in digital infrastructure to support the integration of digital technologies in OH practices. This can include providing access to advanced digital tools, telemedicine platforms, and information systems, that facilitate communication and data sharing between medical and veterinary professionals.

## Data Availability

All the data supporting these findings are found in the results section of the manuscript.

## ACKNOWLEDGEMENT

The authors will like to thank all the students at the University of Buea who participated in the study.

